# Discriminating the prodromal stage of multiple sclerosis using longitudinal health administrative claims data and machine learning–based sequence analysis

**DOI:** 10.1101/2025.09.25.25336630

**Authors:** Ondřej Klempíř, Martina Holá, Martin Rožánek, Juliana Grand Müllerová, Aleš Tichopád

## Abstract

**Background:** Multiple sclerosis (MS) is a chronic autoimmune disease of the central nervous system. Early detection of the prodromal phase could enable timely interventions to potentially modify disease progression. This study leverages longitudinal health administrative claim (HAC) data to identify patterns distinguishing the prodromal stage of MS from other neurological conditions.

**Methods:** HAC data from the Czech Health Insurance Bureau (2017–2022) was analyzed across three cohorts: a target MS cohort with confirmed diagnoses, a control cohort with inconsistent MS suspicions, and a cohort with related disorders. For healthcare utilization and diagnostic code data representation, we employed two approaches: temporal analysis using various time windows relative to the index date (including pre- and post-index date comparisons) and a separate segment-based analysis. Features were extracted using token frequencies and word embeddings. Random forest models were evaluated using Area Under the Receiver Operating Characteristic Curve (AUC) to assess performance.

**Results:** Each cohort included several hundred to over a thousand individuals. The models achieved AUCs around 0.9 for distinguishing the target cohort from controls, with even higher performance in differentiating pre- and post-diagnosis phases. Longer observation windows enhanced predictive accuracy, and feature extraction methods like TF-IDF and word2vec yielded the most consistent results. Segment-based analysis identified a subset of individuals for potential diagnostic reclassification. Interpretable machine learning techniques were integrated into the analysis pipeline.

**Conclusions:** This study highlights the potential of HAC data for detecting early prodromal indicators of MS. Unlike previous research, which often focused on the volume of healthcare utilization, this work explores the informational content within diagnostic codes and healthcare utilization patterns. The findings align with existing research on early neurological condition detection, demonstrating that administrative data could support early identification and intervention in MS and possibly other diseases.

## 1 Background

Analogous to other neurological, psychiatric and neuroimmune disorders, characterizing the multiple sclerosis (MS) prodrome requires extensive research to establish diagnostic criteria [1–5]. Focusing on elucidating the prodromal characteristics and validating high-risk profiles for targeted therapeutics bear potential of fundamentally altering disease prognosis [5, 6]. In addition to clinical markers, healthcare utilization also indicates changes in prodromal MS patients compared to the control group. Exploring the early stages of MS reveals intriguing patterns of increased healthcare interactions preceding formal diagnosis. Research highlights a notable uptick in healthcare usage among MS patients up to five years before their initial demyelinating symptoms become clinically evident [7]. This trend spans various healthcare services, from hospital admissions to prescription pickups, hinting at an underlying prodromal phase, a period marked by subtle, non-specific symptoms that precede the full manifestation of MS. These early signs, while often mirroring common health issues, suggest a critical window for early detection. The challenge lies in distinguishing these early indicators from abundant health complaints being captured in the data too, a task that could potentially be refined by use of algorithms [8–10]. Long-term conditions, including latent viral infections, could potentially be identified through administrative data [11]. The wealth of data from health administrative records presents an untapped resource that could help to timely spot early warning signs [10, 12].

Machine learning methods repeatedly showed to be a robust tool for dissecting administrative healthcare data, allowing for the identification of intricate patterns related to health conditions based on service utilization [13–17]. This technology can uncover subtle signals within the longitudinal data, aiding in early disease detection and optimizing patient care.

The typical administrative medical claim data profile is characterized by a collection of codes, which represent categorical data gathered over time. Given this structure, machine learning models commonly employed in natural language processing (NLP) are well-suited for analysis. Consequently, it is common to conduct categorical encoding to transform these medical codes into numerical values compatible with machine learning algorithms.

Various techniques for NLP-based feature extraction are being utilized in the current literature, such as term frequency-inverse document frequency (TF-IDF), word-to-vector (family of Word2Vec algorithms), or transformer-based methods [18]. Word-to-vector methods and transformers employ dense vectors known as embeddings to encapsulate the meaning of words or sentences. Unlike TF-IDF, where vector elements correspond directly to terms, embeddings lack explicit meanings for individual elements. While embeddings often yield superior performance, they may present challenges in interpretability [19]. On the other hand, generating the significantly simpler TF-IDF representation is typically faster, and the resulting vectors enable the application of inherently interpretable machine learning algorithms, maintaining the interpretability of the resulting models. Recent research indicated that TF-IDF can outperform Word2Vec, particularly in the context of medical records [20]. While new language models like Bidirectional Encoder Representations from Transformers (BERT) exhibit excellent predictive performance on certain tasks, the performance difference between BERT and TF-IDF is sometimes reported to be relatively small, with TF-IDF being an interesting alternative to BERT representation for classification [21].

In recent advanced machine learning-based solutions for claims data, transformer-based language models have been widely applied to improve predictive capabilities. For instance, the Claim Pre-Training (Claim-PT) framework involves training on large-scale claims data, calculating embeddings, and developing models for predictions [22]. Additionally, a transformer-based model has been trained on extensive claims data to predict the progression of severe COVID-19 disease [23]. However, interpreting deep learning models for medical records remains inherently challenging. In terms of the interpretability of transformer-based solutions, a method has been proposed where top words with the most significant influence on identifying specific diseases are extracted using transformer attention mechanisms, with considerable variability in the obtained top words across different models [24].

As previously mentioned, while it is known that patients with MS exhibit increased healthcare utilization prior to their first demyelinating event based on health administrative data, we hypothesized that the content of the recorded information could provide additional discriminative power. Herein, we develop and apply machine learning models to health administrative claims data to identify differences in medical sequences with the goal of classifying and detecting the prodromal phase of MS. To enhance the interpretability of our results, we integrated recently developed interpretable modules into our analysis. Our primary objective was to perform a comprehensive analysis of medical claims records from MS patients and compare them to a control group. As part of this analysis, we aimed to identify recurrent patterns or signals that emerge prior to the diagnosis of MS. These signals may include a wide spectrum of manifestations, ranging from subtle neurological symptoms to nonspecific somatic complaints. Identifying such patterns is important for enhancing diagnostic procedures and facilitating early interventions, potentially mitigating disease progression or alleviating symptoms.

## 2 Methods

### 2.1 Ethical Considerations

The study was approved by the Ethics Committee of the Faculty of Biomedical Engineering (FBME), Czech Technical University in Prague. The authors take full responsibility for all aspects of the work and ensure that any questions regarding the accuracy or integrity of any part of the work will be properly investigated and resolved. All data used in the study is anonymized in accordance with the provisions of HIIPA and GDPR. The activities do not involve linking to other databases or sources that would allow for the identification of patients. The research is exclusively retrospective and non-interventional, conducted within the scope of real-world clinical practice using data generated during routine healthcare. All data processing took place solely within the FBME, and the data is not shared with third parties.

### 2.2 Health administrative claims

Longitudinal health administrative claims (HAC) data consist of codes that uniquely identify specific healthcare utilizations, such as medical examinations, hospital stays, or prescribed medications. These codes are linked to coded medical specialties, offering insights into the expertise and disciplines involved in patient care. Additionally, to some degree diagnosis codes of the International Classification of Diseases, 10th Revision (ICD-10) can reveal the high-level conditions or ailments affecting patients during their healthcare interactions. Detailed measures, stages or conditions such as blood pressure or the Expanded Disability Status Scale (EDSS) are however missing. This makes the HAC data distinctly different from regular electronic health records.

### 2.3 Source database

The Health Insurance Bureau in the Czech Republic provided HAC data from 2017 to 2022. Codes for all utilized health care, including procedures, medications coded according to the Anatomical Therapeutic Chemical (ATC) classification, along with disease codes and specialist codes, were retrieved for each subject, all with corresponding timestamps. The accuracy of this data relies on the correct coding and recording of diagnoses, procedures, and other relevant information in the database.

Out of all data available, the source population included patients treated for G35 (i.e. ICD-10 for MS). The broad patient population was initially defined as individuals aged 18 to 40 who had a recorded instance of the key G35 code. For further refinement into the analytical cohorts, additional codes closely associated with MS were verified. The goal was to establish a cohort of MS patients and a control cohort comprising individuals initially suspected of having MS, but whose diagnosis was not consistently confirmed and was likely replaced by alternative diagnoses over time.

#### 2.3.1 Cohort definition

The Target cohort, likely representing individuals with true MS, was defined by the presence of MS diagnostic codes, the occurrence of magnetic resonance imaging (MRI) codes, and cerebrospinal fluid analysis codes, all recorded for the same individual around the time of diagnosis (determining the index date) or throughout the follow-up period. Additionally, individuals were included if they were repeatedly exposed to MS ICD-10 codes, in combination with codes for neurological specialization and the use of corticosteroids or high-potency MS medications (i.e., Alemtuzumab, Natalizumab, Ocrelizumab, Ofatumumab, Cladribine, Fingolimod, Siponimod, Ozanimod, and Ponesimod). In the target MS cohort, the index date was defined as the date of the first occurrence of the MS code G35 during a visit to a neurologist, following a series of diagnostic visits to various other specialists (Supplementary Table 1).

The series of diagnostic visits comprised the following codes: 29410 (Lumbar or suboccipital cerebrospinal fluid collection), 47357 (Hybrid magnetic resonance imaging and positron emission tomography), 89713 (MR imaging of head, extremities, joints, single segment), and 89725 (Repeat or additional MR imaging - angiography).

In contrast, the Control was identified similarly, except the G35 diagnosis code was inconsistent and eventually discontinued in the records. Moreover, these individuals did not receive any MS-specific treatment, such as corticosteroids or high-potency MS medications, suggesting they did not undergo active treatment for MS. The index date for both the Target and Control cohorts was determined using the same criteria; however, subjects in the Control cohort were not followed up with treatment and were subsequently not considered as having MS (Supplementary Table 1).

The period prior (referred to as the Ante segment or period) and after the index date (referred to as the Post segment or period) were considered separately in the study to facilitate the proposed discrimination analyses.

#### 2.3.2 Related disorders definition

We further, by means of frequency analysis, analyzed patients of the Control cohort for their diagnoses occurring subsequently to identify the most common diagnoses resembling the initially suspected MS. The Control cohort consisted to a great degree of patients with clinically similar diagnoses to MS, but with differences in disease progression and healthcare utilization, which was later reclassified from MS to another disease. Hence, we derived a newly defined set and named it as the Related disorders group. Additional details are provided in Supplementary 1. Subjects in Related disorders do not have a defined index date.

### 2.4 Data preparation

#### 2.4.1 Time windows

To minimize potential selection bias in our machine learning experiments, we avoided imposing strict selection criteria, such as requiring a minimum of one year of prior observation, which could exclude certain patient populations. Instead, we used defined time windows of 360, 180, or 90 days, either before or after the index date, as trimming boundaries. Complete record coverage within the specified time window was not mandatory.

#### 2.4.2 Segments and labeling

Due to the absence of defined onset times (index date) in those Related disorders, making it impossible to delineate pre- and post-periods, segments consisting of 50 codes were obtained and used for classification. For these segments, the diagnosis label was determined by identifying the most frequently occurring individual ICD-10 code (maximum occurrence).

#### 2.4.3 Medical specialization filtering

To account for the large number of codes in the records, we used two types of input sequences. The first, filtered sequence type, included only codes frequently associated with relevant medical specialties, such as neurology, thus highly relevant to MS. The following specialties were found to be highly overrepresented in both the Target and Control cohorts: neurology, standard inpatient neurology, nuclear medicine, radiology, and MRI. Experts confirmed that these specialties are particularly relevant for MS patients.

The second sequence type, referred to as full sequence or the full data, included all codes from the records, even those unrelated to the condition, such as dental visit codes. Since the drugs in the dataset were not linked to specific medical specialties, all medications were included in the analysis without applying any specialty-based filters.

### 2.5 Feature extraction

To model which temporal sequences are likely to correspond to the prodromal phase of MS, we utilized a set of sequence representations commonly employed in NLP. These methods allowed us to capture both token frequency and semantic information from the HAC data. The representations included a baseline model based on the number of observed codes, a count vectorizer, a bag-of-words (BoW) feature vector normalized using TF-IDF, and two semantic word embeddings (word2vec and doc2vec).

The entire feature extraction pipeline was implemented in Python. This process utilized the word2vec package [25], gensim for doc2vec [26] and the scikit-learn sparse matrix representations [27], among other libraries.

#### 2.5.1 Token frequency analysis

For the frequency analysis, we encoded the input sequence as an individual token vector, created a count vectorizer, and generated a TF-IDF matrix. In these approaches, each token was treated as independent, and no relationships between similar words were captured. Each code was considered unique, and its occurrences, reflecting the prevalence of codes in the claims data, were analyzed. For the count vectorizer (cnt-mat), we utilized a unigram model, which served as a frequency-based alternative to one-hot encoding. This method creates a sparse binary matrix by replacing each code with its occurrence count based on its prevalence. In the case of the TF-IDF representation, we implemented a TF-IDF vectorizer, removing terms that appeared too infrequently and excluding terms that were present in fewer than five sequences.

#### 2.5.2 Semantic analysis

For the semantic embedding using word2vec and doc2vec, we trained either word/token-level embeddings (i.e. word2vec) or document-level embeddings (i.e. doc2vec). Each concept was assigned an embedding representation learned through the training of word2vec or doc2vec neural network. The vectors were trained using context, capturing word similarity through the similarity of the vectors themselves. Unlike token frequency analysis, semantic embeddings can handle codes that were not part of the training corpus, providing robust representations even for unseen data.

For the word2vec representation [28], each word is treated as a token and can be trained in a semi-supervised manner using two different strategies: predicting the middle token’s embedding given the surrounding embeddings (continuous bag of words; CBOW) or predicting the surrounding embeddings given the middle embedding (skip-gram). We generated word embeddings by training a 200-dimensional word2vec model, with a window size parameter of 32, and used the skip-gram neural network architecture. This approach is particularly effective at predicting adjacent context words and representing infrequent tokens. The resulting model encoded each token into a numerical vector, which was then processed either by calculating their sum (w2v-sum; sequence length dependent) or mean (w2v-mean; sequence length independent), in order to represent the entire sequence.

For the doc2vec representation, the entire input sequence was considered as a document, and the model was trained at the document level. There are two main variants of the doc2vec approach: distributed bag of words (d2v-dbow), which focuses on how words are distributed within a text, and distributed memory (d2v-dm), which takes into account the context in which words appear. We generated the d2v embeddings by training 768-dimensional models with a window size of 64.

The obtained embedding features were dimensionally reduced using Principal Component Analysis (PCA). The principal components (PCs) were then used in further visualization analysis with the t-distributed Stochastic Neighbor Embedding (t-SNE) method. For the t-SNE method, the perplexity value was empirically set to 30, which successfully separated the cohorts.

### 2.6 Binary classification and evaluation

To evaluate the effectiveness of the proposed data representations in cohorts with a defined index date (Target, Control), we utilized a random forest machine learning model as the primary classification approach. A key advantage of using decision trees within the random forest is their inherent as well as post-hoc interpretability. To address the issue of class imbalance, we implemented a random under-sampling strategy and under-sampled the larger cohort to match the size of the smaller cohort. For robustness and reliability, the dataset was partitioned using a 5-fold cross-validation approach during the main experimentation phase. In this setup, no hyperparameter optimization was performed. Each experiment was repeated five times, and the average model performances were reported for comparison. To evaluate predictive performance, we primarily used the Area Under the Receiver Operator Characteristics Curve (AUC), a widely used metric of discrimination performance that ranges from 0.5 (indicating random prediction) to 1.0 (indicating perfect prediction).

#### 2.6.1 Explainability

To enhance applicability of the presented models and to identify which procedures and drugs had the strongest influence on the model predictions, we implemented the Local Interpretable Model-Agnostic Explanation (LIME) method to explain individual predictions for each model [29]. We assessed the explainability of the random forest models by using TF-IDF features, identifying the most important codes (k = 5) that significantly contributed to the model’s predictions per subject, either positively or negatively.

### 2.7 Multiclass classification

Specific assumptions were made for the multiclass models developed to compare the Target and Control cohorts with the Related disorders. In experiments involving segmented data, with sequences up to 50 codes in length, model training was performed at the patient level. This approach ensured that if a patient’s trajectory consisted of multiple segments, no segment from the same patient was included in both the training and testing datasets. The classifier used for this analysis was the XGBoost classifier, employing multiclass softmax decision-making. To split the available data into training and testing sets, we utilized the default train-test function from the scikit-learn package [27], with a division of 75% for the training set and 25% for the test set. A multiclass confusion matrix was reported to evaluate and interpret performances across various conditions.

## 3 Results

### 3.1 Cohort characteristics

The total number of patients for Target, Control, and Related disorders groups were 743, 1161, and 969, respectively, with 30.1% males and 69.9% females in the Target group, 29.7% males and 70.3% females in the Control group, and 40.8% males and 59.2% females in the Related disorders group. The mean ages were 30.21, 31.36, and 31.42 years, respectively. The Related disorders group comprises patients with the most abundant diagnoses codes as given in Table 1.

**Table 1.** Distribution of subjects in Related disorders.

| ICD-10 code | Condition | Sample size | Percentage (%) |
| --- | --- | --- | --- |
| G98 | Other disorders of the nervous system, not elsewhere classified | 190 | 19.6 |
| R42 | Dizziness and giddiness | 118 | 12.2 |
| R202 | Paresthesia of skin<br>(abnormal sensations like tingling or numbness) | 112 | 11.6 |
| H46 | Optic neuritis<br>(inflammation of the optic nerve) | 76 | 7.8 |
| R51 | Headache | 102 | 10.5 |
| G939 | Disorder of the brain, unspecified | 70 | 7.2 |
| G379 | Demyelinating disease of the central nervous system, unspecified | 301 | 31.1 |
| **Total number of patients** |  | **969** | **100** |

The table (Table 2) presents the number of reported diagnosis codes for the defined Target and Control cohort in detail. The distribution of code counts when divided into the periods before and after the index date revealed significant variability in the accumulated number of codes across individual subjects.

**Table 2.** Quantity of codes reported for each cohort (full data; sequences not filtered).

|  | Total |  | Ante |  | Post |  |
| --- | --- | --- | --- | --- | --- | --- |
|  | Target | Control | Target | Control | Target | Control |
| **Sample size** | 743 | 1161 | 676 | 1085 | 743 | 1161 |
| **Mean** | 713 | 573 | 154 | 192 | 572 | 394 |
| **Median** | 616 | 484 | 114 | 143 | 467 | 304 |
| **Min** | 103 | 59 | 1 | 1 | 21 | 12 |
| **Max** | 3187 | 5137 | 1093 | 1314 | 3187 | 4218 |
| **Unique specialties** | 183 | 190 | 135 | 158 | 172 | 178 |

### 3.2 Capability to discriminate Target vs. Control

The model’s ability to discriminate between the Target and Control cohorts was evaluated by calculating the AUCs on the filtered data across three pre- and post-diagnosis windows: 90, 180, and 360 days.

For the Control cohort (n = 1161), within the 180-day pre-post window, we analyzed the distribution of the number of codes per subject, finding a mean of 50 codes (std: 28, min: 3, max: 266). By setting a threshold of 25 as the minimum number of codes required for a subject (i.e. sequence) to be included in the model training and evaluation, the sample size was reduced to 982 subjects, with an updated mean of 55 codes (std: 26). Applying the same threshold to the Target cohort (743 subjects, with a mean of 61 codes, standard deviation: 27, min: 2, max: 205) resulted in 700 subjects being included in the model, with an updated mean of 63 codes (std: 25). This threshold for the minimum number of codes was also applied to the 90-day and 360-day pre-post windows, leading to the exclusion of some subjects and consequently altering the sample sizes across these windows.

#### 3.2.1 Classification

Overall, the results demonstrated that model performance improved with longer observation windows, indicating that additional data allowed for more accurate predictions (Table 3). Among the methods evaluated, TF-IDF and cnt-mat showed the highest overall performance (AUC ~ 0.9), particularly in the longest window (360 days), closely followed by the word2vec and doc2vec methods. As the length of the pre-post window increased, we observed a corresponding increase in the AUC for the model trained on the number of codes. However, this baseline classifier remained a weak predictor when compared to the other representation methods (AUC ~ 0.6).

**Table 3.** The cross-validated AUC performance metrics.

| pre-post window | 90 days | 180 days | 360 days |
| --- | --- | --- | --- |
| *(balanced; downsampled)* | (N = 1216) | (N = 1400) | (N = 1462) |
| # codes | 0.54 | 0.58 | 0.62 |
| cnt-mat | 0.81 | 0.88 | 0.90 |
| TF-IDF | 0.82 | 0.88 | 0.91 |
| w2v-sum | 0.77 | 0.84 | 0.87 |
| w2v-mean | 0.77 | 0.84 | 0.86 |
| d2v-dbow | 0.73 | 0.80 | 0.85 |
| d2v-dm | 0.75 | 0.77 | 0.76 |

The table (Table 3) presents the cross-validated AUC performance metrics for various data representation methods across different pre-post windows (90, 180, and 360 days). The performance of each method is reported for increasing lengths of the observation window, with sample size (N) provided for each window. The results indicate that model performance generally improves with longer observation windows, and with frequency analysis methods, focusing on occurrences of the individual codes, showing the highest overall performance.

#### 3.2.2 Explainability experiment

By means of the LIME method, we extracted the two lists representing the top 10 conditions or procedures reported for Target vs Control, reflecting significantly different expressed patterns in healthcare utilization between these groups (Suppl. Table 2). Overall, the first list (Target) seemed to reflect a cohort that is more focused on ongoing treatment and management of a chronic condition, with several therapeutic interventions and less emphasis on diagnostic evaluations (medications like “Copaxone”, “Methylprednisolon,” and “Interferon beta-1a” indicate specific therapeutic interventions). The second list (Control) was more oriented towards diagnostic and follow-up evaluations, suggesting a cohort that may be under active monitoring for neurological conditions. LIME evaluation was conducted on all available subjects. Selected example sequences from both Target and Control cohorts, showing the most contributing codes, are visualized in Fig. 1.

**Fig. 1.**
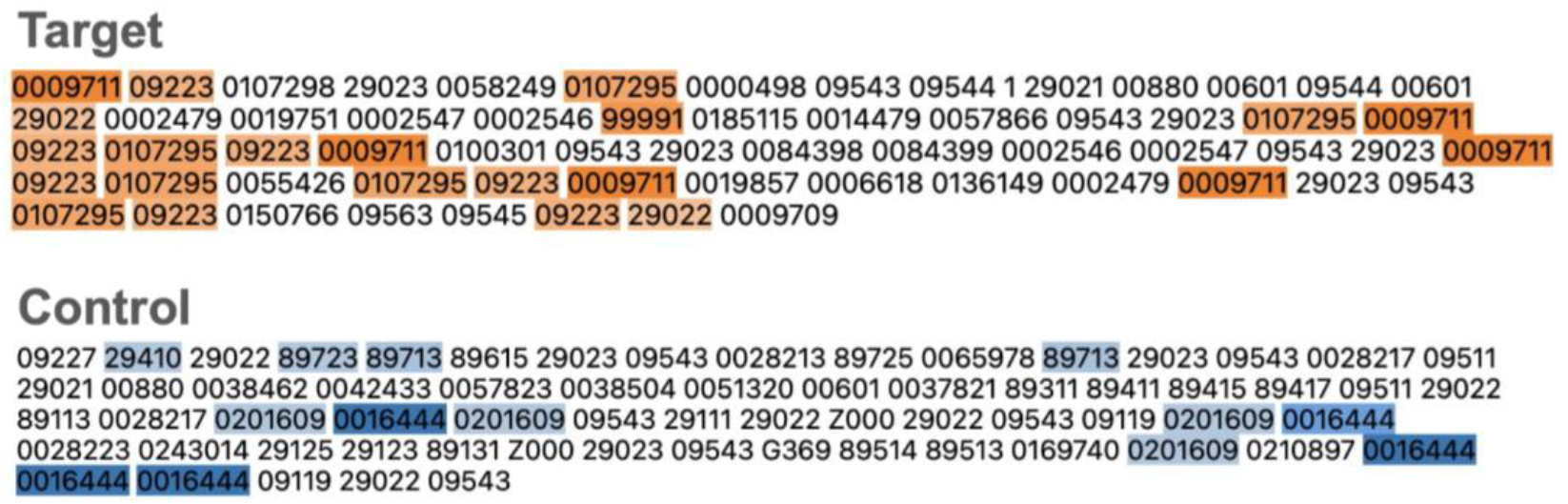
Example sequences with high code importance obtained by LIME Explainer. The LIME method extracted the codes that significantly influence the model’s prediction decisions. For the Target cohort (orange) and Control cohort (blue), the top 5 most contributing codes for the given cohort prediction are illustrated. In the Control cohort, the presence of carbamazepine (N03AF01), an antiepileptic drug, is shown, as well as electrophysiological examinations like EEG (29113, 29115, 29123, 29125, 29140, 29145, 29150, 91714) and EMG (29210, 29220, 29230, 29240). On the other hand, methylprednisolone (H02AB04) is typical for the Target cohort. It is evident that the frequency of certain codes can have a greater impact than individual possibly important but rare codes. This characteristic aligns with the nature of the TF-IDF classifier, where the frequency of tokens is shown to have a greater impact than the inherently important events, which typically occur only once within the analysis window (180 days before and after the index date in this case). Individual codes can be mapped back to a human readable form and annotated using the annotation lists available at [30].

### 3.3 Capability to discriminate pre-index vs. follow-up phase

Next, we evaluated the model’s effectiveness in predicting the pre-index (prodromal) versus follow-up phase (Table 4) and assessed the effect of filtering.

**Table 4.** Data representation methods in predicting the class (pre vs. post index) under different scenarios.

|  | Control Cohort |  | Target Cohort |  |
| --- | --- | --- | --- | --- |
|  | Filtered | Full data | Filtered | Full data |
|  | (N = 432) | (N = 1666) | (N = 294) | (N = 1012) |
| # codes | 0.53 | 0.62 | 0.72 | 0.73 |
| cnt-mat | 0.90 | 0.91 | 0.96 | 0.97 |
| TF-IDF | 0.90 | 0.91 | 0.95 | 0.97 |
| w2v-sum | 0.85 | 0.86 | 0.94 | 0.95 |
| **w2v-mean** | 0.85 | 0.86 | 0.91 | 0.94 |
| **d2v-dbow** | 0.77 | 0.86 | 0.77 | 0.94 |
| **d2v-dm** | 0.77 | 0.76 | 0.83 | 0.85 |

For the Control filtered cohort, within 180 days before onset, we started with 1008 subjects, having an average of 17 codes (standard deviation of 15, ranging from 1 to 158). After applying a minimal length threshold, the dataset was reduced to 216 subjects, with an increased average of 39 codes (standard deviation of 17). In the 180 days after onset, the initial 1,161 subjects had an average of 35 codes (standard deviation of 22, with a range of 2 to 157). Post-threshold, the dataset consisted of 740 subjects, with an average of 46 codes (standard deviation of 20).

For the Target filtered cohort, within 180 days before onset, 607 subjects were analyzed, with an average of 17 codes (standard deviation of 14, ranging from 1 to 97). After applying the minimal length threshold, this was reduced to 147 subjects, with an average of 37 codes (standard deviation of 11). In the 180 days after onset, the initial cohort of 743 subjects had an average of 47 codes (standard deviation of 23, with a range of 2 to 202). After thresholding, 644 subjects remained, with an average of 52 codes (standard deviation of 21).

For the Control full sequence cohort, in the 180 days before onset, we began with 1076 subjects, averaging 72 codes (standard deviation of 68, ranging from 1 to 772). After thresholding, 833 subjects were retained, with an average of 90 codes (standard deviation of 68). For the 180 days after onset, the initial 1161 subjects averaged 121 codes (standard deviation of 75, with a range of 2 to 653). After applying the minimal length threshold, the dataset was reduced to 1122 subjects, with an average of 125 codes (standard deviation of 74).

In the Target full sequence cohort, 665 subjects in the 180 days before onset averaged 66 codes (standard deviation of 51, with a range of 1 to 262). After thresholding, this was reduced to 506 subjects, with an average of 83 codes (standard deviation of 47). In the 180 days after onset, the initial cohort of 743 subjects averaged 145 codes (standard deviation of 78, ranging from 2 to 650). After applying the minimal length threshold, 727 subjects remained, with an average of 148 codes (standard deviation of 76).

#### 3.3.1 Classification

For the Control cohort, both filtered and full sequence scenarios show that the use of data representation techniques like TF-IDF and word/document embeddings significantly improve the predictive performance compared to simply using the number of codes. In the filtered sequence scenario, the AUC values for TF-IDF and w2v models reach 0.90, while the baseline model (#codes) performs considerably lower at 0.53. This pattern holds in the full sequence scenario, with TF-IDF and w2v methods achieving higher AUC values of 0.91 and 0.86 respectively, again surpassing the baseline.

For the Target cohort, the filtered sequence scenario shows even higher AUC values across all methods compared to the Control cohort. The TF-IDF method and word embeddings (w2v-sum, w2v-mean) perform exceptionally well, reaching AUC values as high as 0.96 and 0.95 respectively. The full sequence scenario follows a similar trend with slightly improved AUC values across all methods. The doc2vec models (d2v-dbow, d2v-dm) perform better in the full sequence scenario, indicating that they may capture important document-level semantic information when more data is available.

Data representation methods in predicting the class (pre vs. post index) under different scenarios (Table 4), as measured by the cross-validated AUC values. Also, the performance of each method is reported for the effect of filtering, with sample size (N) provided for each subcohort. The results indicated that model performances (except for the #codes baseline model) achieved very high AUC scores and on top of that, AUCs did not drop on the full sequence data.

#### 3.3.2 Explainability experiment

To demonstrate the effect of filtering on identifying the key medical codes in the prodromal phase (180 days before the index date) for the Target cohort, we analyzed the top codes that contributed positively towards identifying the prodrome. The interpretable lists of relevant medical concepts generated for this phase are documented in Supplementary Table 3. This evaluation was conducted using the LIME method on all available subjects within the specified scenario.

The comparison of the two lists of top conditions in the prodromal phase revealed distinct characteristics between the filtered and full data. As expected, the first list, which applied neurologically relevant filtering, primarily included conditions and procedures related to neurological assessments. These included MRI imaging of the head and limbs, lumbar puncture for cerebrospinal fluid collection, and various electrodiagnostic tests (e.g. EMG and EEG). The presence of mephenoxalone, a muscle relaxant, also indicates a focus on neurological or musculoskeletal conditions. In contrast, the second list, which included all medical specialties without filtering, was dominated by common diagnostic tests and general medical evaluations. These included blood tests (prothrombin test, cholesterol, triacylglycerols), CT scans, and of high interest, ophthalmologic procedures. The inclusion of targeted eye examinations suggests that vision-related issues were significant in the prodromal phase within this cohort.

We discovered that “noise” in the data can also contribute valuable insights, for example, it revealed important biochemical tests that are genuinely associated with an MS diagnosis, making them relevant. These tests, though routine, became important due to their frequency, which contributed to their relevance. Moreover, a significant finding in the full data was the emphasis on eye care, which underscores the broader range of specialties captured without filtering. The explainability analysis on full data demonstrated that the method could help identify additional relevant specialties, suggesting its utility even in the presence of noise.

### 3.4 Discriminating prodromal trajectories vs. Related disorders

Ultimately, we focused on predicting individual segments and distinguishing the MS prodromal phase from other neurological conditions. For the Related disorders group, where index date data was unavailable, we opted to work with filtered data, segmenting each patient’s journey into sequences of 50 codes. This segmentation generally captured the entire patient journey for the filtered data. The final diagnosis for each segment was determined based on the most frequently occurring diagnosis code within that sequence.

#### 3.4.1 Examining segment-level similarities and dissimilarities

We hypothesized that subjects with the same trajectory or disease are grouped together, while patients that are diagnosed with different diseases are well-separated. This observation indicates that our framework is able to capture meaningful clinical information within claims data and learning efficient embeddings (Fig. 2).

**Fig. 2.**
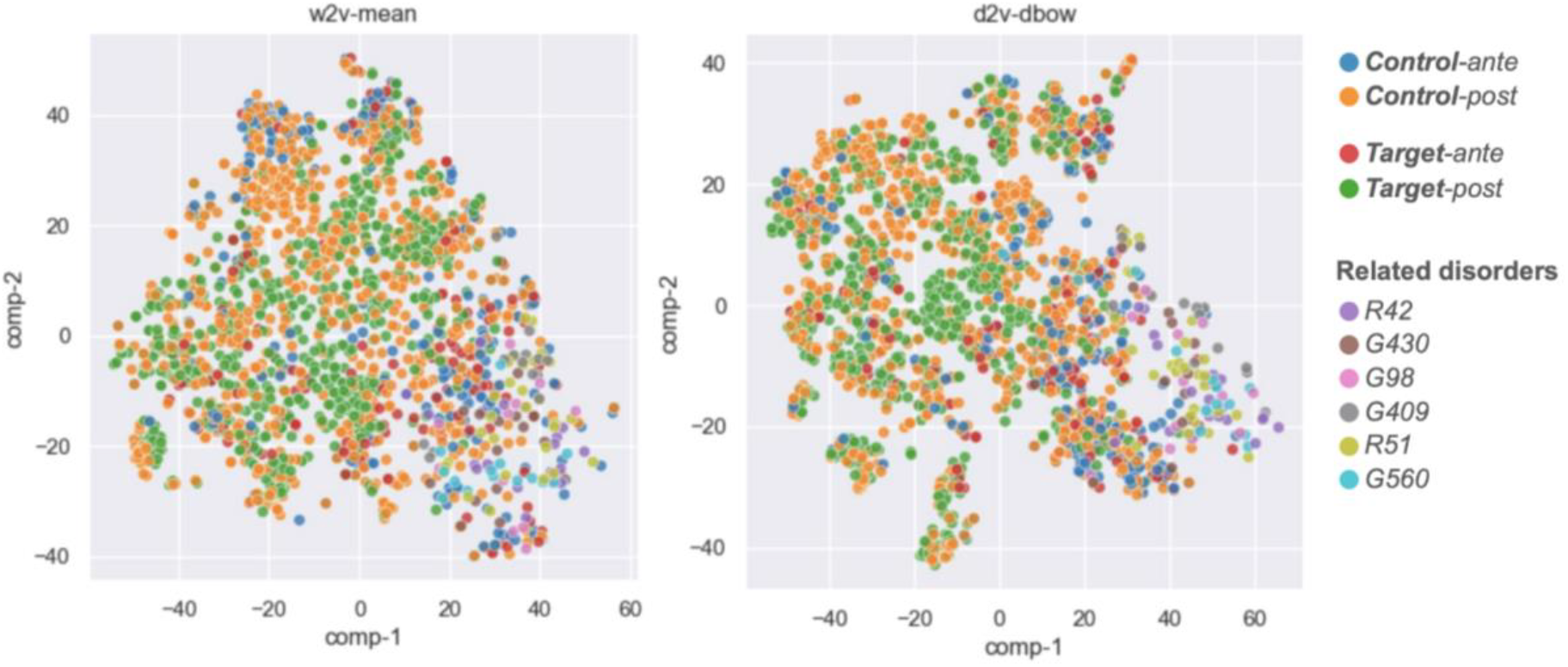
Embeddings visualization. T-SNE scatter plots of word2vec-mean (left) and doc2vec-dbow (right) learned. It is evident that Ante segments (red, blue) are typically positioned closer to the top 6 most common ICD-10 diseases from the Related disorders group (i.e., R42: Dizziness and Giddiness, G430: Migraine without aura, G98: Other disorders of nervous system, G409: Epilepsy, R51: Headache, G560: Carpal tunnel syndrome). ICD-10 max. occurrence diagnosis that describes the corresponding disease condition was determined for each segment presented.

#### 3.4.2 Binary classification

Herein, we further hypothesized that a portion of segments with relevant neurological disorders from the Related disorders group could potentially be reclassified as part of the MS (Target) prodromal phase. To test this hypothesis, we implemented a binary classifier using w2v-mean embedding. We trained a two-class balanced XGBoost classifier on filtered 50-length segments from the MS (Target) prodromal cohort (positive class; N = 147) and 50-length segments from the Related disorders group (negative class; available N = 373), using a total of 294 samples with a 75/25% train/test split.

By evaluating the confusion matrix on the test set, we achieved a classification accuracy of 93%, with 4% of subjects identified for potential reclassification (marked as False Positives). When we expanded the testing to include the entire dataset, the accuracy improved to 96%, with 4% of segments suggested for reclassification.

As part of the additional testing, we also included an evaluation on the Control-ante segments (N = 216), a portion of data not used for model training but hypothesized to have similar symptoms and events as Target prodromal, though not followed up as MS. This external test run classified 78% of Control-ante as Target-ante, with only 22% assigned to the Related disorders.

#### 3.4.3 Multiclass classification

To obtain a comprehensive view of the model’s ability to classify segments into different classes and to distinguish the Related disorders group from all other segments, we implemented a multiclass classification scenario (Fig. 3).

**Fig. 3.**
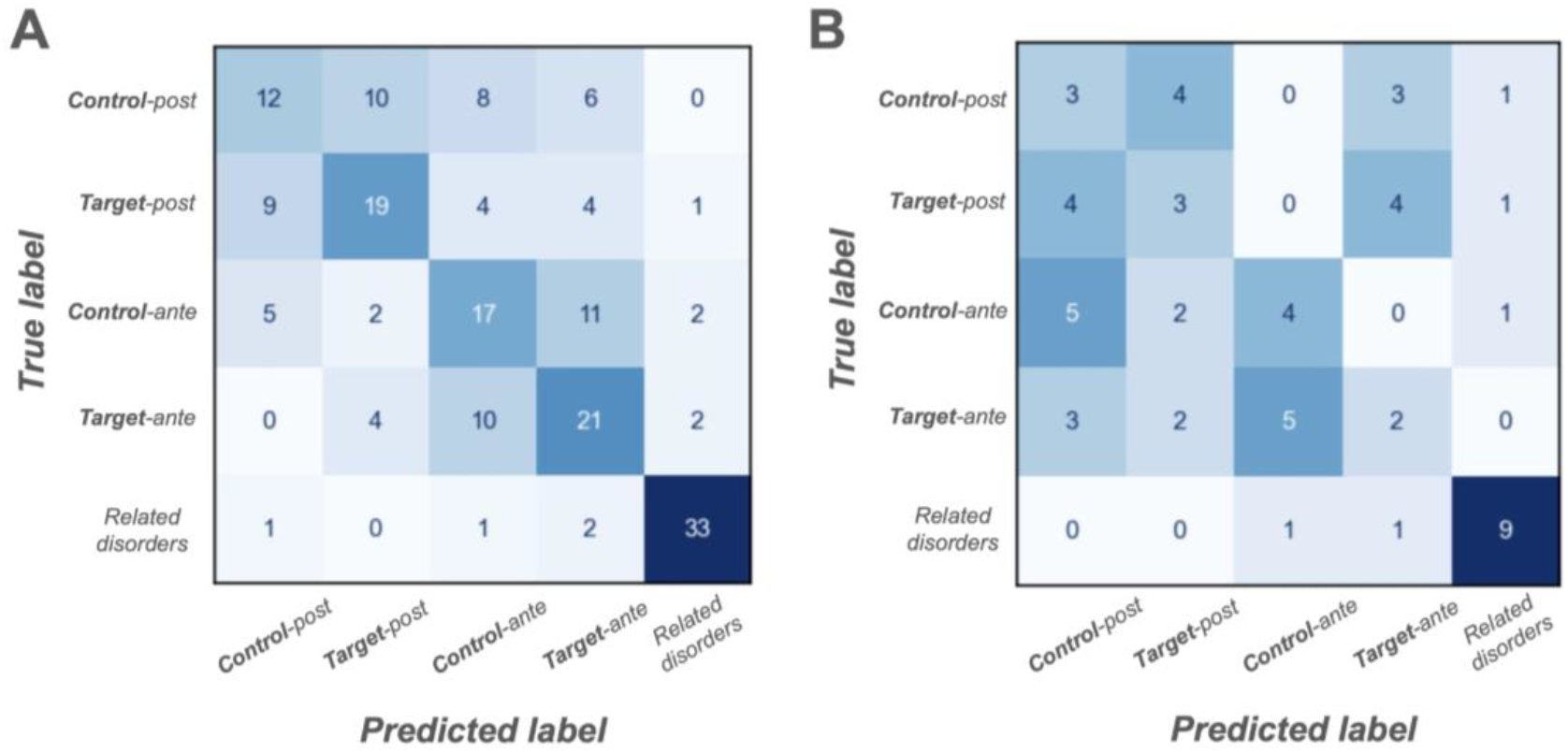
The multiclass classification results are presented via two confusion matrices. For the w2v-mean model on the test sets: A) Target and Control segments exhibit similarities in their definitions, leading to higher misclassification rates, particularly between Ante and Post phases within the same cohort. Despite this, the model demonstrates a strong ability to distinguish Target-like segments from the Related disorders. B) When ATC medications were removed from the sequences, the ability to differentiate between Target and Control decreased, reflecting a drop in classification performance. However, the distinction between Target-like segments and the Related disorders group remained largely unaffected.

## 4 Discussion

This article offers insights into the prodromal phase of multiple sclerosis through the examination of health administrative data and machine learning techniques. The main findings of our study demonstrate the ability to retrospectively identify the prodromal phase of MS using the patients’ observational medical records. This approach has the potential to facilitate early intervention and enhance long-term prognosis.

In the last decade, population-based studies have emerged that have employed objective methods to examine symptoms occurring prior to the onset of MS [5, 7, 31–37]. The collective findings of these studies indicate that the prodromal phase of MS can be identified a minimum of five years prior to the onset of MS symptoms or up to a maximum of ten years before the initial diagnosis of the disease. These individuals tend to visit healthcare providers more frequently for non-neurological issues, such as anxiety or depression. Additionally, they often undergo MRI scans due to headaches, which, while neurological in nature, may reveal findings consistent with MS, specifically radiologically isolated syndrome (RIS). RIS represents a radiological manifestation of MS without overt MS symptoms. All of this information can be extracted from administrative data. In individuals with RIS, the prodromal phase has been observed to have a variable duration, potentially commencing as early as ten to fifteen years before the manifestation of MS symptoms [38, 39].

Recent studies have increasingly focused on the utilization of administrative health data to identify the prodromal phase of various diseases, including MS. This approach employs the analysis of large datasets, including insurance claims and medical records, to identify patterns and early symptoms that precede a formal diagnosis. The utilization of this data is of particular value in the study of neurological diseases, such as MS and Parkinson’s disease (PD) [40–42]. The aforementioned patterns, when analyzed retrospectively, may facilitate the construction of predictive models to identify individuals at elevated risk of developing MS before the onset of typical symptoms [42, 43]. Similar approaches have been employed in PD, where healthcare claims data has been utilized to examine the prevalence of early symptoms and patterns that may correspond to prodromal stage [40].

One of the primary contributions of this study is the high degree of discriminative ability obtained by our machine learning models to effectively discriminate between MS patients and controls. Models based on TF-IDF and embedding methods exhibited high AUC values (higher than 0.9). To discuss our findings, we developed three distinct contrasts to model different scenarios, each providing a different form of insights into distinguishing the prodromal phase of MS. “Model 1” (Target vs. Control) showed that while the Target and Control cohorts share similarities, our models were able to differentiate them effectively, indicating the presence of subtle but meaningful differences. “Model 2” (Pre-vs. Post-diagnosis) demonstrated strong discriminative performance within both the Target and Control cohorts, regardless of whether filtering by medical specialties was applied, highlighting the robustness of this approach. “Model 3” (Segment classification), which incorporated all three cohorts (Target, Control, and Related disorders) in a single analysis, confirmed the observed similarity between the Target and Control groups while clearly distinguishing them from the more dissimilar Related disorders group.

Our analysis demonstrated that the proposed machine learning-based system can help with identifying candidates for potential reclassification. In our case, this included flagging approximately 4% of individuals from the Related disorders cohort as being in the prodromal phase of MS. These cases may represent instances of misdiagnoses or missed opportunities for earlier recognition of MS. Quantitatively, the 4% can be extrapolated to the incidence or prevalence of MS. In the Czech Republic, approximately 700–1000 patients initiate their first disease-modifying therapy each year during the follow-up period [44]. Based on this, our system could identify several dozen individuals annually who might benefit from earlier reclassification and initiation of treatment. It is important to note, however, that some of these individuals may have eventually been classified as MS patients through standard diagnostic pathways. The value of this approach lies in accelerating the diagnosis process. Nonetheless, such a system would require practical validation and long-term monitoring to assess its effectiveness and ensure its reliability in real-world clinical settings.

A further significant outcome of our investigation was the capability of the LIME (Local Interpretable Model-agnostic Explanations) approach to elucidate concepts and codes that informed the models’ decision-making processes. As demonstrated by Rajkomar et al. (2019) [45], it is crucial to comprehend the rationale behind model predictions to ensure their acceptance and credibility in healthcare settings.

We explored various time windows and filtering strategies to optimize the detection of the prodromal phase of MS. For scenarios with known index dates, we employed fixed time periods before and after diagnosis. In the segment-based approach, we analyzed sequences in fixed segments of 50 codes. Additionally, filtering by medical specialization allowed us to narrow the scope of codes to neurologically relevant trajectories without losing informational value compared to classifications performed on full sequences. Furthermore, the results suggested that focusing on health code frequencies (TF-IDF) may be more effective than incorporating broader contextual information (doc2vec).

A comparison of our results with those of other studies examining the prodromal phase of neurological diseases reveals a trend in the detection of non-specific symptoms preceding the onset of disease manifestations. For example, studies from 2017 [7] and 2022 [5] assert the significance of early detection of MS through the analysis of health data, while also confirming that MS patients demonstrate an elevated rate of healthcare utilization several years prior to diagnosis. Similarly, our findings demonstrate that machine learning can discern significant patterns in administrative data that are characteristic of the prodromal phase of MS. Our findings are consistent with those of Ravaut et al. (2021) [17], who also employed machine learning to predict health conditions based on administrative data and achieved comparable success in predicting diabetes-related complications. Additionally, a study by Delpino et al. (2022) [46] effectively employed machine learning to anticipate the onset of chronic illnesses based on electronic health records, thereby corroborating the assertion that machine learning can forecast the prevalence of specific chronic ailments, their progression, their underlying causes, and in numerous scenarios. Another study [47] demonstrated the efficacy of machine learning in predicting cardiovascular complications based on electronic health records.

While the results of our study are promising, it is important to consider the limitations. One limitation of the study is the restricted sample size, which was limited to approximately 1000 subjects. While the sample size was sufficient to examine the prodromal phase of MS, larger cohorts could enhance the robustness and generalisability of the results. Although some transformer models can already be trained on large-scale claims data, comprising millions of samples, as in the present study [22], at later stage, i.e. for downstream tasks, approximately 500 cases are selected and 1000 controls (negatives) are chosen, which is a very similar sample size to that in the present study. Moreover, it has been demonstrated that even large pretrained models require further fine-tuning. A further limitation of the study is that the time interval between events is not addressed. Further investigation and more detailed analyses may be inspired by the study conducted by Lentzen [23].

Although our results are promising, future research should include more sophisticated machine learning techniques to further enhance our capacity to identify the prodromal phase of MS and, consequently, yield more precise, primarily clinical, conclusions.

## 5 Conclusion

It is well-established that healthcare utilization increases prior to multiple sclerosis diagnosis. Our primary hypothesis was whether machine learning leveraging the informational content of utilization could enhance predictive power. We focused on various machine learning scenarios, such as Target vs. Control and Pre-vs. Post-diagnosis.

The findings of our study indicated that the analysis of administrative health data through the utilization of advanced machine learning methodologies can serve as an efficacious instrument for the identification of the prodromal phase of multiple sclerosis. The models based on TF-IDF and embedding methods exhibited a high degree of discriminatory power between MS patients and controls. In terms of predictive capability, compared to traditional statistical evaluations based solely on the volume of codes and utilization, our system can identify patients who may be suitable for reclassification. To enhance the interpretability of our results, we integrated recently developed interpretable modules into our analysis. This highlights its potential utility in improving early detection and intervention strategies.

Although the present study was focused on MS disease, it is believed that the approach developed has the potential to be applied to other diseases, which could significantly contribute to improving diagnosis and treatment in a broader context.

## Data availability

Access to de-identified CZ HAC patient data is restricted to researchers affiliated with FBME and is not available for public sharing. Additional supporting data and code required to reproduce the results will be made available upon reasonable request.

## Abbreviations

ATC: Anatomical Therapeutic Chemical
AUC: Area Under the Receiver Operating Characteristic Curve
BERT: Bidirectional Encoder Representations from Transformers
CBOW: continuous bag of words
Claim PT: Claim Pre-Training
cnt-mat: Count vectorizer
CT: Computed Tomography
d2v-dbow: distributed bag of words
d2v-dm: distributed memory
EDSS: Expanded Disability Status Scale
EEG: Electroencephalography
EMG: Electromyography
HAC: Health administrative claim
ICD-10: International Classification of Diseases, 10th Revision
LIME: Local Interpretable Model-Agnostic Explanation
MRI: Magnetic resonance imaging
MS: Multiple Sclerosis
NLP: Natural language processing
PCA: Principal Component Analysis
PCs: Principal components
PD: Parkinson’s disease
TF-IDF: Term frequency-inverse document frequency
t-SNE: Stochastic Neighbor Embedding
Word2Vec: word-to-vector

## Ethics approval and consent to participate

This study complies with the Declaration of Helsinki and has also been approved by the Ethics Committee of the Faculty of Biomedical Engineering, Czech Technical University in Prague. The authors take full responsibility for all aspects of the work and ensure that any questions regarding the accuracy or integrity of any part of the work will be properly investigated and resolved. All data used in the study are anonymized in accordance with the provisions of HIIPA and GDPR. The activities do not involve linking to other databases or sources that would allow for the identification of patients. The research is exclusively retrospective and non-interventional, conducted within the scope of real-world clinical practice using data generated during routine healthcare. All data processing takes place solely within the Faculty of Biomedical Engineering, and the data is not shared with third parties. Given the nature of the data and the approval obtained from the Ethics Committee of the Faculty of Biomedical Engineering, Czech Technical University in Prague, informed consent from patients was not required.

## Contributors

Conceptualization: OK, MH, MR, JM, AT; Methodology: OK, MH, MR, JM, AT; Interpretation: OK, MH, JM, AT; Writing-first draft: OK, MH, AT; Writing-review and editing: OK, MH, JM, AT; Funding acquisition: MR.

## Acknowledgements

We extend our sincere gratitude to all involved parties whose support and collaboration were instrumental in the successful completion of this project.

This research was funded by the following grants: the project CZ.02.01.01/00/23_025/0008743, funded by the European Union under the Operational Programme Johannes Amos Comenius (OPJAK); OPTAK MPO89679/24/61400; and SGS24/153/OHK5/3T/17.

## References

1. Meisenzahl E, Wege N, Stegmüller V, Schulte-Körne G, Greimel E, Dannlowski U, et al. Clinical high risk state of major depressive episodes: Assessment of prodromal phase, its occurrence, duration and symptom patterns by the instrument the DEpression Early Prediction-INventory (DEEP-IN). Journal of Affective Disorders. 2024;351:403–13.

2. Mellergaard C, Waldemar G, Vogel A, Frederiksen KS. Characterising the prodromal phase in dementia with Lewy bodies. Parkinsonism & Related Disorders. 2023;107:105279.

3. Yao L, Liang W, Chen J, Wang Q, Huang X. Constipation in Parkinson’s Disease: A Systematic Review and Meta-Analysis. Eur Neurol. 2023;86:34–44.

4. Hawkes CH. The prodromal phase of sporadic Parkinson’s disease: Does it exist and if so how long is it? Movement Disorders. 2008;23:1799–807.

5. Marrie RA, Allegretta M, Barcellos LF, Bebo B, Calabresi PA, Correale J, et al. From the prodromal stage of multiple sclerosis to disease prevention. Nat Rev Neurol. 2022;18:559–72.

6. Roman S. A journey with no roadmap—The need for validated criteria of the MS prodrome. Mult Scler. 2023;29:502–4.

7. Wijnands JMA, Kingwell E, Zhu F, Zhao Y, Högg T, Stadnyk K, et al. Health-care use before a first demyelinating event suggestive of a multiple sclerosis prodrome: a matched cohort study. The Lancet Neurology. 2017;16:445–51.

8. Benchimol EI, Guttmann A, Mack DR, Nguyen GC, Marshall JK, Gregor JC, et al. Validation of international algorithms to identify adults with inflammatory bowel disease in health administrative data from Ontario, Canada. Journal of Clinical Epidemiology. 2014;67:887–96.

9. Schuler KP, Hemnes AR, Annis J, Farber-Eger E, Lowery BD, Halliday SJ, et al. An algorithm to identify cases of pulmonary arterial hypertension from the electronic medical record. Respir Res. 2022;23:138.

10. Gillmeyer K, Nunez E, Rinne S, Qian S, Klings E, Wiener R. DEVELOPMENT AND VALIDATION OF ALGORITHMS TO IDENTIFY PULMONARY ARTERIAL HYPERTENSION IN ADMINISTRATIVE DATA. Chest. 2020;158:A2421–2.

11. Antoniou T, Zagorski B, Loutfy MR, Strike C, Glazier RH. Validation of Case-Finding Algorithms Derived from Administrative Data for Identifying Adults Living with Human Immunodeficiency Virus Infection. PLoS ONE. 2011;6:e21748.

12. Mathai SC, Hemnes AR, Manaker S, Anguiano RH, Dean BB, Saundankar V, et al. Identifying Patients with Pulmonary Arterial Hypertension Using Administrative Claims Algorithms. Annals ATS. 2019;16:797–806.

13. Campbell RL, Alpern ML, Li JT, Hagan JB, Motosue M, Mullan AF, et al. Development of a machine learning algorithm based on administrative claims data for identification of ED anaphylaxis patient visits. Journal of Allergy and Clinical Immunology: Global. 2023;2:61–8.

14. Ravaut M, Harish V, Sadeghi H, Leung KK, Volkovs M, Kornas K, et al. Development and Validation of a Machine Learning Model Using Administrative Health Data to Predict Onset of Type 2 Diabetes. JAMA Netw Open. 2021;4:e2111315.

15. Van Deynse H, Cools W, De Deken V-J, Depreitere B, Hubloue I, Kimpe E, et al. Predicting return to work after traumatic brain injury using machine learning and administrative data. International Journal of Medical Informatics. 2023;178:105201.

16. Sharma V, Kulkarni V, Mcalister F, Eurich D, Keshwani S, Simpson SH, et al. Predicting 30-Day Readmissions in Patients With Heart Failure Using Administrative Data: A Machine Learning Approach. Journal of Cardiac Failure. 2022;28:710–22.

17. Ravaut M, Sadeghi H, Leung KK, Volkovs M, Kornas K, Harish V, et al. Predicting adverse outcomes due to diabetes complications with machine learning using administrative health data. npj Digit Med. 2021;4:24.

18. Al-Fuqaha’a S, Al-Madi N, Hammo B. A robust classification approach to enhance clinic identification from Arabic health text. Neural Comput & Applic. 2024;36:7161–85.

19. Zarsky J, Lopez G, Kliegr T. Explainability of Text Clustering Visualizations—Twitter Disinformation Case Study. IEEE Comput Grap Appl. 2022;42:8–19.

20. Seinen TM, Kors JA, van Mulligen EM, Fridgeirsson EA, Verhamme KM, Rijnbeek PR. Using clinical text to refine unspecific condition codes in Dutch general practitioner EHR data. International Journal of Medical Informatics. 2024;189:105506.

21. Piskorski J, Haneczok J, Jacquet G. New Benchmark Corpus and Models for Fine-grained Event Classification: To BERT or not to BERT? In: Proceedings of the 28th International Conference on Computational Linguistics. Barcelona, Spain (Online): International Committee on Computational Linguistics; 2020. p. 6663–78.

22. Zeng X, Linwood SL, Liu C. Pretrained transformer framework on pediatric claims data for population specific tasks. Sci Rep. 2022;12:3651.

23. Lentzen M, Linden T, Veeranki S, Madan S, Kramer D, Leodolter W, et al. A Transformer-Based Model Trained on Large Scale Claims Data for Prediction of Severe COVID-19 Disease Progression. IEEE J Biomed Health Inform. 2023;27:4548–58.

24. Luo X, Gandhi P, Zhang Z, Shao W, Han Z, Chandrasekaran V, et al. Applying interpretable deep learning models to identify chronic cough patients using EHR data. Computer Methods and Programs in Biomedicine. 2021;210:106395.

25. word2vec · PyPI. 2024. https://pypi.org/project/word2vec/. Accessed 17 Aug 2024.

26. Gensim: topic modelling for humans. 2024. https://radimrehurek.com/gensim/models/doc2vec.html. Accessed 17 Aug 2024.

27. scikit-learn: machine learning in Python — scikit-learn 1.5.1 documentation. 2024. https://scikit-learn.org/stable/. Accessed 17 Aug 2024.

28. Mikolov T, Chen K, Corrado G, Dean J. Efficient Estimation of Word Representations in Vector Space. 2013.

29. Ribeiro MT, Singh S, Guestrin C. “Why Should I Trust You?”: Explaining the Predictions of Any Classifier. 2016.

30. Zdravotní výkony - Zdravotní výkony. 2024. https://szv.mzcr.cz/. Accessed 17 Aug 2024.

31. Yusuf FLA, Ng BC, Wijnands JMA, Kingwell E, Marrie RA, Tremlett H. A systematic review of morbidities suggestive of the multiple sclerosis prodrome. Expert Review of Neurotherapeutics. 2020;20:799–819.

32. Yusuf FL, Wijnands JM, Kingwell E, Zhu F, Evans C, Fisk JD, et al. Fatigue, sleep disorders, anaemia and pain in the multiple sclerosis prodrome. Mult Scler. 2021;27:290–302.

33. Wijnands JM, Zhu F, Kingwell E, Zhao Y, Ekuma O, Lu X, et al. Five years before multiple sclerosis onset: Phenotyping the prodrome. Mult Scler. 2019;25:1092–101.

34. Högg T, Wijnands JMA, Kingwell E, Zhu F, Lu X, Evans C, et al. Mining healthcare data for markers of the multiple sclerosis prodrome. Multiple Sclerosis and Related Disorders. 2018;25:232–40.

35. Wijnands JMA, Zhu F, Kingwell E, Zhao Y, Evans C, Fisk JD, et al. Prodrome in relapsing-remitting and primary progressive multiple sclerosis. Euro J of Neurology. 2019;26:1032–6.

36. Disanto G, Zecca C, MacLachlan S, Sacco R, Handunnetthi L, Meier UC, et al. Prodromal symptoms of multiple sclerosis in primary care. Annals of Neurology. 2018;83:1162–73.

37. Cortese M, Riise T, Bjørnevik K, Bhan A, Farbu E, Grytten N, et al. Preclinical disease activity in multiple sclerosis: A prospective study of cognitive performance prior to first symptom. Annals of Neurology. 2016;80:616– 24.

38. Lebrun-Frenay C, Kantarci O, Siva A, Sormani MP, Pelletier D, Okuda DT, et al. Radiologically Isolated Syndrome: 10-YEAR Risk Estimate of a Clinical Event. Annals of Neurology. 2020;88:407–17.

39. Okuda DT, Siva A, Kantarci O, Inglese M, Katz I, Tutuncu M, et al. Radiologically Isolated Syndrome: 5-Year Risk for an Initial Clinical Event. PLoS ONE. 2014;9:e90509.

40. Warden MN, Searles Nielsen S, Camacho-Soto A, Garnett R, Racette BA. A comparison of prediction approaches for identifying prodromal Parkinson disease. PLoS ONE. 2021;16:e0256592.

41. Tremlett H, Marrie RA. The multiple sclerosis prodrome: Emerging evidence, challenges, and opportunities. Mult Scler. 2021;27:6–12.

42. Tremlett H, Munger KL, Makhani N. The Multiple Sclerosis Prodrome: Evidence to Action. Front Neurol. 2022;12:761408.

43. Makhani N, Tremlett H. The multiple sclerosis prodrome. Nat Rev Neurol. 2021;17:515–21.

44. Stastna D, Drahota J, Lauer M, Mazouchova A, Menkyova I, Adamkova J, et al. The Czech National MS Registry (ReMuS): Data trends in multiple sclerosis patients whose first disease-modifying therapies were initiated from 2013 to 2021. Biomed Pap Med Fac Univ Palacky Olomouc Czech Repub. 2024;168:262–70.

45. Rajkomar A, Dean J, Kohane I. Machine Learning in Medicine. N Engl J Med. 2019;380:1347–58.

46. Delpino FM, Costa ÂK, Farias SR, Chiavegatto Filho ADP, Arcêncio RA, Nunes BP. Machine learning for predicting chronic diseases: a systematic review. Public Health. 2022;205:14–25.

47. Dalal S, Goel P, Onyema EM, Alharbi A, Mahmoud A, Algarni MA, et al. Application of Machine Learning for Cardiovascular Disease Risk Prediction. Computational Intelligence and Neuroscience. 2023;2023:9418666.

