## Supplementary Materials for "Discriminating the prodromal stage of multiple sclerosis using longitudinal health administrative claims data and machine learning–based sequence analysis"

### Supplementary material

S. Table 1. Data definition in cohorts.

| | Target | Control | Related disorders |
|---|---|---|---|
| **Diagnosis code(s)** | G35 | G35 | G98, R42, R202, H46, R51, G939, G379 |
| **Examination codes** | 47357, 89713, 89725, 89723, 29410 | 47357, 89713, 89725, 89723, 29410 | |
| **Expertise code** | 209 | 209 | |
| **Drug codes** | H02AB04, H02AB02, L01XC02, L03AB07, L03AB08, L03AB13, L03AX13, L04AA23, L04AA27, L04AA31, L04AA34, L04AA36, L04AA40, L04AA42, L04AA50, L04AC07, L04AX07, N07XX09, L01FA02, L01XC10, L04AA52. | | |

S. Table 2. Target vs. Control: TOP10 the most abundant codes predicted by LIME. Presenting filtered data, time window 180 days before and after index date.

| **Contributing positively (tended to Target)** | | **Contributing negatively (tended to Control)** | |
|---|---|---|---|
| **99991** | Code only for centres according to Decree 368/2006 - used for reporting ZULP applied outside the centre or outpatient ZULP applied in the centre (together with clinical examination) | **29410** | Collection of cerebrospinal fluid by lumbar or suboccipital puncture or by puncture through the fontanelle |
| **00880** | Resolution of reported hospitalisation = new hospitalisation | **0009552** | Nonspecific |
| **09223** | Intravenous infusion in an adult or child over 10 years of age | **89713** | MR imaging of the head, limbs, joints, one section of the spine (C, Th, or L) |
| **1000000** | Nonspecific | **09543** | Signal performance of clinical examination / until 31.12.2014: regulatory fee per visit - fee paid |
| **0012023** | Cholecalciferol | **29021** | Comprehensive examination by a neurologist |

| | | | |
|---|---|---|---|
| **0214739** | Copaxone | **29123** | Follow-up examination by a neurologist |
| **0009711** | Methylprednisolon | **29210** | EMG examination of nerve conduction velocity |
| **09220** | Peripheral vein cannulation including infusion<br><br>Procedures performed on an outpatient basis aggregated into a treatment day | **29111** | Special neurological examination tests |
| **09543** | Signal performance of clinical examination / until 31.12.2014: regulatory fee per visit - fee paid | **89725** | Repeated or additional MRI scans |
| **0500512** | Interferon beta-1a | **Z000** | General medical examination (check-up) |

S. Table 3: Target filtered (left) | Target full sequence (right) - TOP10 codes. Contributing positively (towards Ante).

| **Contributing positively (tended to Ante)** | | **Contributing positively (tended to Ante)** | |
|---|---|---|---|
| **89713** | MR imaging of the head, limbs, joints, one section of the spine (C, Th, or L) | **96623** | Prothrombin test |
| **29410** | Collection of cerebrospinal fluid by lumbar or suboccipital puncture or by puncture through the fontanelle | **89615** | CT scan with a larger number of scans (over 30), without the use of contrast medium |
| **89725** | Repeated or additional MRI scans | **96163** | Blood count |
| **1000000** | Nonspecific | **75161** | Non-contact tonometry (1 eye) |
| **29210** | EMG examination of nerve conduction velocity | **81471** | Cholesterol - total |
| **29125** | EEG using activation methods - evaluation | **09563** | Performance of emergency medical services |

| | | | |
|---|---|---|---|
| **29123** | EEG using activation methods (technical version only) | **81611** | Triacylglycerols |
| **29220** | EMG examination of reflexes, neuromuscular transmission and tetany | **75022** | Targeted examination by an ophthalmologist |
| **0085656** | Mefenoxalon | **75163** | Refraction examination with autorefractor (1 eye) |
| **89615** | CT scan with a larger number of scans (over 30), without the use of contrast medium | **Z000** | General medical examination (check-up) |
